# Health literacy of inland population in the mitigation phase 3.2. of COVID-19’s pandemic in Portugal - a descriptive cohort study

**DOI:** 10.1101/2020.05.11.20098061

**Authors:** Joana Gomes da Silva, Carla Sofia Silva, Bárbara Alexandre, Pedro Morgado

**Author notes:** Correspondence: Rua Santa Catarina, n.° 6, 5370-407, Mirandela, Portugal.

## Abstract

**Background:** COVID-19 is a respiratory disease transmitted through respiratory droplets with a high transmission rate. There’s still no effective and approved antiretroviral treatment or vaccine, thus, preventive measures are the main key to contain this pandemic. Portugal is now in phase 3.2 of the mitigation of COVID-19, being imperative to understand the health literacy of our country and how to prevent the lack of information, through community empowerment.

**Material and methods:** A cross-sectional study with a cohort from a primary care facility was conducted. We collected demographic and epidemiological data and carried out a questionnaire by phone call. Descriptive statistics and nonparametric tests were used to assess associations between the independent variables and the level of health literacy. The level of significance was set at p<0.05.

**Results:** Our cohort includes 222 subjects (median age: 62 years old), mostly females (131), undergraduate (193) and with at least one risk factor for COVID-19 (144). Globally, younger individuals, females, graduates and the Non-Risk Group appear to have higher levels of health literacy, with some exceptions to this pattern.

**Conclusions:** We observe a well-informed population. However, being a pandemic situation, we intend to identify and correct outliers/misconceptions. This work allows a perspective of the current state/pattern of health literacy as well as its possible predictors. Furthermore, this study makes aware of how relevant the specific communication approaches are. Further studies to understand the predictors of health literacy are necessary.

## 1. INTRODUCTION

In December 2019, an outbreak of pneumonia with unknown etiology began to arise in the city of Wuhan, China and the diagnosis of influenza and other Coronaviruses (such as MERS and SARS) were considered but later excluded by laboratory tests^1,2^. On 7^th^ of January, 2020, China announced that the cause of that epidemic was a new Coronavirus strain, later designated SARS-CoV-2 and proven to be present in wild bats^2,3,4^.

The transmission mode of SARS-nCoV-2 is through respiratory droplets, and they are either a source of direct or indirect contamination^5,6^. This strain presented a basic reproduction number (R0) within a range from 1.4 to 6.5: the highest infection rates in the elderly or in people with underlying pathologies^2,7^. Therefore, this means that this strain of virus has a high rate of human-to-human transmission and a person is much more contagious as more symptomatic^6,8^.

The median age of the symptomatic patients seems to be between 47-59 years even though it can affect individuals of all ages^2^.

The clinical presentation of COVID-19 has a wide spectrum that ranges from assymptomatic or slightly upper respiratory infection to septic shock^6,9,10^.

The cardinal symptoms described by the World Health Organization and later integrated in the flowcharts issued by the Direção Geral de Saúde (DGS) are “cough”, “fever” and “dyspnea”. However, other studies report myalgia, fatigue, headache and gastrointestinal symptoms as other symptoms to be taken into account^11,12,14^. In a study involving 1099 laboratory confirmed cases, Zhong *et al* described that the most common clinical manifestations were fever (88.7%), cough (67.8%) and fatigue (38.1%), with sputum, dyspnea, sore throat and headache also being highly reported by the patients^13^. This study also mentions that there was a minor percentage of patients who showed gastrointestinal symptoms such as vomiting and diarrhea.

Despite the broad spectrum of the disease, Huang *et al* described that the elderly or those with underlying pathologies such as hypertension, Chronic Obstructive Pulmonary Disease, Diabetes Mellitus and cardiovascular diseases had a faster progression of the disease, with a higher rate of Acute Respiratory Distress Syndrome, consequent multiorgan failure and higher rate of death^2,10^. These pathologies, considered as risk factors for COVID-19, were later included in the “Norma de Orientação Clínica 004/2020” of DGS, as patients with a particular risk of infection and with higher care needs.

At the moment, there is no effective and approved antiretroviral treatment or vaccine targeting SARS-CoV-2, with the treatment being mainly symptomatic and organ support^6,10^.

Therefore, in the absence of effective specific treatment, there is a range of preventive measures to be taken, such as a correct handwashing, respiratory etiquette, disinfection of surfaces and social isolation and/or social distancing (> 2 meters)^2,3,6,14^. These measures are assumed as vital in the control of the mitigation of the pandemic^3,14^.

To raise public awareness of these measures and health promotion, community empowerment is necessary and the main key for this public health problem. This empowerment through health literacy programs and official campaigns using television, radio and other media has a positive impact, especially in this actual context - outbreaks and/or pandemics^15-17^.

On the 18^th^ of March, 2020, the State of Emergency was decreed in Portugal, with a Law Decree regulating preventive measures to contain the pandemic mitigation in Portugal. On the 26^th^ of March, 2020, phase 3.2 of the mitigation of COVID-19’s pandemic in Portugal was declared, e.g., chains of community transmission of the virus in the national territory. Thereupon, it arose the need for tighter control and application of preventive measures.

Regarding the positive impact of official campaigns and community empowerment with a strong health literacy, we considered it extremely important to study the health literacy of the population in this mitigation phase, in order to understand the flaws that may still exist as well as identifying possible predictors of health literacy concerning this matter. Thus, and since as far as we’re concerned this is the first study focusing on this subject, it will be possible to rectify possible wrong ideas through specific intervention strategies aimed at different populations. Also, we hope that this study helps to identify possible errors/flaws of health literacy in a pandemic situation and to avoid them at the early beginning of a possible future pandemic.

## 2. MATERIAL AND METHODS

### Subjects and Data Collection

We conducted this cross-sectional study with a cohort of 222 subjects, with an age greater or equal to 15 years old - to avoid a misunderstanding bias of our questionnaire - COVID-19’s Questionnaire (Supplementary Information).

We excluded subjects codified with the ICP-2 codes for “dementia”, “mental retardation” or “presbycusis”, - to exclude the inherent misunderstanding bias.

We selected a cohort: individuals with a scheduled medical appointment in a primary care facility of Unidade Local de Saúde de Nordeste between 1st and 8th of April.

We collected the demographic and epidemiological data (age, gender, education level and risk factor(s) for COVID-19 codified by ICP-2) and carried out COVID-19’s Questionnaire by phone call with simultaneous registration of the data, between March 30 and April 3.

### Statistical Analysis

For statistical analysis, we regarded “Age” as a continuous variable. Concerning the other variables, we categorized subjects according to their “Gender” (“Female” and “Male”); Education (“Undergraduate Group” - <4 years of schooling; 4, 6, 9 or 12 years of schooling; “Graduate Group” - Graduate, Master’s Degree and/or Doctorate); “Risk factor(s) for COVID-19 codified by ICP-2” (“Risk Group” - presence - and “Non-risk Group” - absence - as described in “Norma de Orientação Clínica 004/2020” by Direção Geral de Saúde (DGS) such as Chronic Obstructive Pulmonary Disease, Asthma, Cardiac Insufficiency, Diabetes Mellitus, Chronic Liver Disease, Chronic Renal Disease, Ative Malignant Neoplasm or Immunosuppression State).

We performed the statistical analysis of the collected data using Microsoft Office Excel 2019® (Microsoft, Redmond, Washington, USA) and the SPSS® statistical package (standard version 22.0; SPSS, Chicago, IL, USA). Exploratory analysis was conducted to demographically characterize our cohort as well as for the answers given for each question of our questionnaire.

We used non-parametric tests (Kruskal Wallis H and Fisher’s Exact Test) to test whether significant associations between the variables and the answers given were observed or not, except for the last question.

The level of significance for all statistical tests was set at p<0.05, with a 95% confidence interval.

This study was submitted for approval and approved by the Direction of Department of Primary Health Care of Unidade Local de Saúde do Nordeste, according to the Declaration of Helsinki of the World Medical Association. The confidentiality of the data was guaranteed, only accessible by the main investigator and the respective authors.

## 3. RESULTS

### 3.1. Sample characterization

This study comprises a cohort of 222 subjects, with an age range 15-94 years old and a median age of 62 years old. The majority registered an undergraduate level of education and had at least one risk factor for COVID-19 codified by ICP-2 (Table 1).

**Table 1.** Demographic characterization of cohort.

|  |  | **Absolute Frequency** | **Relative Frequency (%)** | **Median age (years)** |
| --- | --- | --- | --- | --- |
| **Sex** | **Females** | **131** | 59.01 | 62 |
|  | **Males** | **91** | 40.99 | 61 |
| **Education** | **Undergraduate Group** | 193 | 86.94 |  |
|  | **Graduate Group** | 29 | 13.06 |  |
| **Risk Factor(s) for COVID-19 codified by ICP-2** | Non-Risk Group | 78 | 35.14 | 49 |
|  | Risk Group | 144 | 64.86 | 66 |

### 3.2. COVID-19’s Questionnaire

Globally, younger individuals, females, graduates and the Non-Risk Group presented higher relative frequencies of the correct answer along COVID-19’s Questionnaire. However, three exceptions were observed: the Undergraduate Group and the Risk-Group had a high relative frequency stating that COVID-19 has a cure and in mentioning “Social Isolation” as an important preventive measure to adopt when compared to the Graduate Group and the Non-Risk Group, respectively. Males have higher relative frequency in answering the correct number of SNS24 and in stating that children can get sick and transmit the infection by SARS-CoV-2 when compared to females (Table 2 - Supplementary information).

The use of Non-parametric tests (Kruskal Wallis H and Fisher’s Exact Test) demonstrated several statistically significant associations between our variables and the answers given along COVID-19’s Questionnaire (Table 3).

**Table 3.** Nonparametric tests applied for the correct answers throughout COVID-19’s Questionnaire.

| **Variables** | **Age** |  | **Gender** | **Education** | **Risk factor** |
| --- | --- | --- | --- | --- | --- |
| **Correlation coefficient/ Association test** | **Point-biserial correlation coefficient** | **Kruskal Wallis H Test** | **Fisher's Exact Test** |  |  |
|  |  | **p-value** | **p-value** | **p-value** | **p-value** |
| Question 1:<br>Answer "Yes" | -0.20 | 0.26 | 0.70 | 1.00 | 0.09 |
| Question 2:<br>Answer "Other" | -0.22 | 0.10 | 0.67 | <0.01* | 0.06 |
| Question 2:<br>Answer "Fever" | -0.23 | 0.28 | 0.31 | 0.20 | 0.09 |
| Question 2:<br>Answer "Cough" | -0.1 | 0.06 | 0.22 | 0.65 | <0.01* |
| Question 2:<br>Answer "Dyspnea" | 0.03 | 0.52 | 1.00 | 0.14 | 0.37 |
| Question 3:<br>Answer "Cure" | - | 0.53 | 0.78 | 0.02* | 0.01* |
| Question 4:<br>Answer "Other" | -0.14 | 0.20 | 0.59 | 1.00 | 0.01* |
| Question 4:<br>Answer "Respiratory Etiquette" | -0.02 | 0.84 | 0.15 | 1.00 | 0.46 |
| Question 4:<br>Answer "Handwashing" | -0.10 | 0.85 | 0.27 | 0.16 | 0.12 |
| Question 4:<br>Answer "Social isolation" | 0.04 | 0.61 | 0.41 | <0.01* | <0.01* |
| Question 5:<br>Answer "Stay home and call SNS 24" | -0.26 | 0.02* | 0.04* | 1.00 | 0.03* |
| Question 6:<br>Answer "808242424" | -0.18 | 0.54 | 0.04* | 0.49 | 0.03* |
| Question 7:<br>Answer "No" | - | 0.84 | 0.13 | 1.00 | 0.09 |
| Question 8:<br>Answer "No" | - | 0.70 | 0.04* | 0.81 | 0.34 |
| Question 9:<br>Answer "No" | - | 0.25 | 0.01* | 0.09 | <0.01* |
| Question 1:<br>Answer "No" | - | 0.48 | 0.74 | 0.40 | <0.01* |
| Question 11: Answer "Yes" | - | 0.88 | 0.35 | 0.19 | 0.22 |
| Question 12:<br>Answer "Yes" | - | 0.92 | 0.32 | 0.16 | 0.53 |

\*p-value set at 0.05. Statistically significant differences observed.

#### 3.2.1. QUESTIONS REGARDING THE COURSE OF COVID-19

When questioned about the symptomatology of COVID-19, 96.85% of individuals stated that they know the symptoms of COVID-19. Indeed, analysing only the 215 individuals who answered “Yes” in the first question, we observe that most of the individuals stated the cardinal symptoms of COVID-19 described in several “Normas de Orientação Clínica” from DGS (e.g. “Fever”, “Cough” and “Dyspnea”) in relative frequencies >70.00% and that 36.28% stated “Other” symptoms such as “myalgia”, “headache” and “loss of smell and taste”. Nonparametric tests denote statistically significant associations: “Other” reveal a significant association with “Education”, with individuals of Graduate Group stating more often other symptoms of the disease; “Cough” reveal a significant association with “Risk Factor”, with individuals from Non-Risk Group stating more often this symptom.

When questioned whether COVID-19 only affects old people or not, 90.99% of individuals answered the correct answer. Nonparametric tests reveal a statistically significant association regarding variable “Gender”, with females answering more often the correct answer.

Regarding the two questions about children, there’s a good level of health literacy among our cohort. When questioned if children can get sick, there’s a major dominance of the answer “Yes” (88.74%). Nonparametric tests reveal a statistically significant association regarding variable “Risk Factor”, with individuals from “Non-Risk Group” stating more often the correct answer.

When we questioned about the possibility of children transmitting the disease, 83.78% of individuals answered “Yes”. No statistically significant associations were observed.

Regarding the question about the cure of COVID-19, we found a great dispersion of the answers: 41.44% of the subjects refer that COVID-19 has a cure, 33.33% deny it and 25.23% refer that they don’t know whereas this disease has a cure or not. Nonparametric tests reveal a statistically significant association regarding variable “Education” and “Risk Factor”, with individuals from “Undergraduate Group” and “Risk Group” stating more often the correct answer.

#### 3.2.2. PROCEDURE IN CASE OF SUSPECT OF COVID-19 AND PREVENTIVE MEASURES

When questioned about the correct procedure in case they have symptoms compatible with COVID-19, 77.48% stated they should “Stay home and call SNS 24”. Nonparametric tests reveal a statistically significant association regarding variable “Age”, “Gender” and “Risk Factor”, with younger individuals, females and individuals from Risk-Group stating more often the correct answer.

Regarding the number of SNS 24, only 24.77% of subjects knew the correct number. Nonparametric tests reveal a statistically significant association regarding variable “Gender” and “Risk Factor”, with males and individuals from Non-Risk Group answering the correct number.

When questioned about the preventive measures to adopt, 77.93% of individuals stated “Social Isolation”, 50.90% stated “Other” preventive measures (e.g., the use of gloves, the use of mask, distance >2 meters from other people, leaving shoes at front door,…), 45.50% of individuals refer “Washing hands” and lastly, only 8 subjects (3.60%) of all subjects mention “Respiratory Etiquette” as an important measure to adopt. Nonparametric tests demonstrate several statistically significant associations: “Other” has a statistically significant association with variable “Risk Factor” - individuals from “Non-Risk Group” state this symptom more often - and “Social Isolation” has a statistically significant association with “Education” and “Risk Factor” - individuals from “Graduate Group” and “Risk Group” state this preventive measure more often.

Regarding the use of gloves, a great dispersion of answers was observed, with 56.31% of individuals referring that the use of gloves does not always prevent the infection by the new Coronavirus, 13.51% affirming “I don’t know” and 30.18% answering “Yes”. Nonparametric tests demonstrate that variables “Gender” and “Risk Factor” have a statistically significant association with the answers given, with females and individuals from “Non-Risk Group” answering correctly more often.

Regarding the use of masks, a great dispersion of answers was observed, with 59.91% of individuals referring that the use of gloves does not always prevent the infection by the new Coronavirus, 15.32% affirming “I don’t know” and 24.77% answering “Yes”. Nonparametric tests show a statistically significant association between variables “Risk Factor” for this question, with individuals from “Non-Risk Group” answering correctly more often.

When we questioned our subjects about the possibility of visits of friends and/or family during social isolation, 83.33% stated “No”, 12.16% stated “Yes” and 4.5% affirmed “I don’t know”. Nonparametric tests show no statistically significant associations.

#### 3.2.3. SOURCE OF INFORMATION

Regarding the final question, and even though we have a slight dispersion of frequencies of answers, it is obvious that the major source of information about COVID-19 is the television, with 74.77% of individuals reporting that fact. After television, the social networks (10.36%) and other (4.50%) - such as official websites (WHO, DGS and CDC), employers, Town Hall and Guarda Nacional Republicana - have an important role in informing the population about the actual public health problem (Fig. 1).

**Figure 1.**
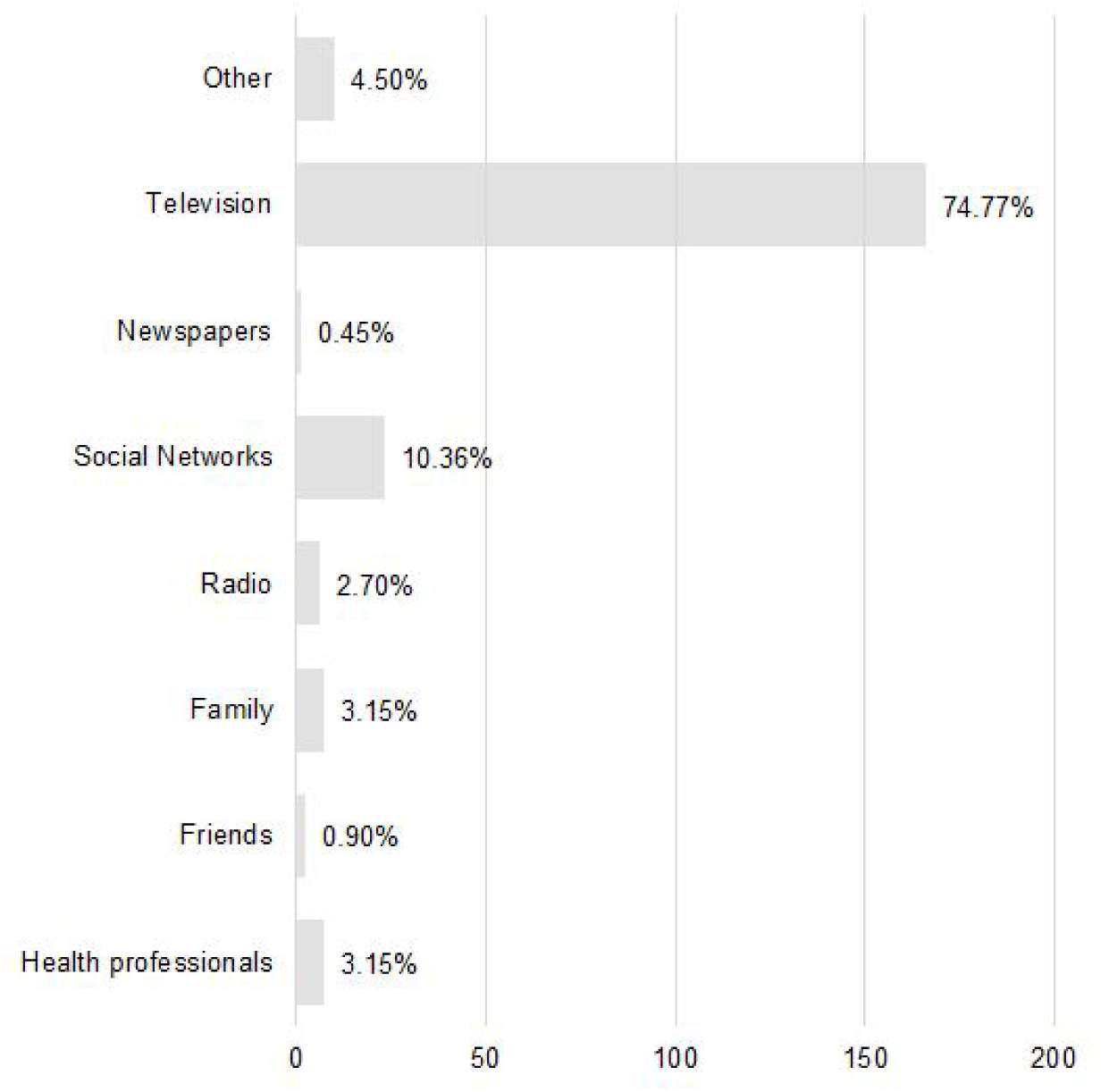
Absolute and relative frequencies of answers for the fourteenth question.

## 4. DISCUSSION

According to Portugal’s censuses, the population of our cohort comprises 23850 citizens, with a proportion >1 female per male and as well as an undergraduate and older population^18,19^. The characterization of our cohort is compatible with these data.

Globally, our results demonstrate a relatively well informed population. Furthermore, females, younger individuals, the “Graduate” and the Non-Risk Groups presented higher relative frequencies of the correct answer throughout our questionnaire. There are some exceptions to this pattern, which we are going to discuss later on this section.

In the literature, several studies support our results, reporting age, gender and education as predictors of health literacy. Indeed, Sørensen *et al* in their comparative study of the results of European Health Literacy survey (HLS-EU) reported education, age and gender as predictors of low health literacy, along with social status and financial privation. According to their findings, males and older individuals tend to have slightly lower health literacy, while a higher level of education is a strong positive predictor of health literacy^20^. Besides, regarding health literacy, several other studies state these gender, age and education significant differences, reporting that an increase in age, being male and lower levels of education usually implies a decrease in health literacy^21-27^.

Regarding the presence/absence of risk factors for COVID-19 as far as we’re concerned, there’s no literature available to support the results found in our study. However, we do know that the median age of the Risk Group is higher than the median age of the Non-Risk Group and these differences stated in non-parametric tests along this study may be a consequence of an age bias.

Concerning the symptomatology, most of the population knows the cardinal symptoms of COVID-19 related by DGS (high relative frequencies >70.00%). Curiously, even though not being reported by DGS, there’s a significant percentage of subjects relating. “Other” symptoms such as “myalgia”, “headache” and “loss of taste and smell”, described in several articles^10,12,13,14,28^. This fact may suggest that people do not obtain their information by a single source of information and that they try to search for relevant information.

The third question was thus constructed to capture the discord underlying the definition of “cure”. According to Portuguese lexicon, “cure” is defined as “act or effect of self-healing or healing somebody” or “health recovery” and “healing” being described as “restore health”. However, the answers given along the phone calls highlighted the uncertainty in this definition, emphasizing the insecurity in answering this question. Indeed, throughout this study, most of the people who answered “No”, justified using sentences such as “there’s no vaccine” or “there’s not a medication targeting the virus”. These findings underline some incongruences and misconceptions. A vaccine is a tertiary preventive measure administered in an individual to confer immunity to a certain infectious disease, which enables an asymptomatic/mild clinical manifestation of it, useful in controlling epidemics^29-31^. Therefore, it is necessary to demystify the idea that a vaccine is a cure for an infectious disease. Even though there’s not a specific antiretroviral targeting SARS-CoV-2 efficiently approved, there are several reported cases of healed patients, matching with the Portuguese definition of “cure”^6,10,28,32^. Interestingly, the Undergraduate Group presents a higher relative frequency of the answer “Yes”, yet, it wasn’t possible to find any information in literature supporting this finding.

Regarding the preventive measures to adopt face to COVID-19, overall, the subjects in our study were able to mention them, with the exception of Respiratory Etiquette. In the literature there are several studies that report the importance of handwashing, respiratory etiquette and social isolation in containing epidemics and in controlling SARS-CoV-2 transmission^28,33^. Indeed, even though Chao Yang reports that there’s no proven efficacy of handwashing in controlling SARS-CoV-2 transmission^34^, there are several findings that report the opposite. Qing-Xia *et al*, in a study involving 7 countries, alert to the fact that handwashing significantly slows the exponential spread of SARS-CoV-2^35^ and several other studies showed that a correct handwashing is useful in controlling epidemics as well as SARS transmission^21,35,36^. Even though handwashing is an important preventive measure to adopt face to COVID-19, unfortunately, there are findings that show that the correct procedure - as recommended by WHO - isn’t always applied and that some mistakes are observed, as such low frequency in washing hands and not washing hands the enough time^37,38^. Also, Zhang *et al* conducted a study in Beijing’s population as a post-pandemic assessment and verified that even though people knew the importance of handwashing, they did not apply it^14^. In their study, Fung *et al* also observed a decline of practice of this preventive measure^14^. Therefore, in order to avoid this decline and errors, we must implement methods to successfully achieve a correct preventive handwashing^39^.

As well as for other respiratory viruses, respiratory etiquette has a preponderant role in controlling the transmission of an infectious respiratory disease^40-42^. Looking at the numbers in our study, the absolute and relative frequencies of subjects who mentioned it as a necessary preventive measure was disappointingly low. Thus, this must be a point to improve in health literacy.

Concerning “Social Isolation”, being SARS-CoV-2 an extremely high contagious virus, an effective and important preventive measure to adopt and to contain the pandemic is social isolation^6,43^. Interestingly and contrary to the other questions, in this question the Risk Group has a higher relative frequency of the preventive measure “Social Isolation” than the Non-Risk Group. This fact may be explained by the fact that Non-Risk Group is a younger group. Thus, younger people belong to the proletariat more than older people, having the necessity to leave home for work, which can be a justification to this finding. Regarding the answers obtained in question 7, there are still cases of people who believe it is possible to receive or visit family and/or friends at home. Since this idea may promote the creation of new transmission chains, it is necessary to demystify it, across every single outlier.

Regarding the right procedure in case of having symptoms, most of the subjects stated that they must stay at home and call SNS 24, as recommended by DGS. Concerning the number of SNS 24, interestingly, males report higher relative frequency of the right answer. This may be explained by the fact that in our population, males have higher rates of education with more educational opportunities^19^. Recalling that the Graduate Group has a higher relative frequency of correct answers, this difference between females and males may be explained by this bias. Also, along the phone calls, individuals that belong to the Risk Group - overall, older individuals - referred that they didn’t know the number by heart because their caregivers knew or they had it pointed on a paper. Besides suggesting the lower capacity of older people to memorize numbers, this draws our attention for the important role that caregivers may have in transmitting the proper information to older people.

Regarding the questions about the epidemiology of COVID-19, there’s a major consensus that this disease affects the elderly and that children either transmit and can get sick. Indeed, concerning all the information already mentioned, the literature states that SARS-CoV-2 may infect individuals of all age ranges, with a more severe clinical manifestation in the elderly and/or people with underlying pathologies^6,10,44^. Also, concerning the clinical manifestation and transmission in children, throughout this study, some people reported that “children can transmit the virus, that’s why grandchildren and grandparents cannot be together”. Indeed, children can get sick and are an important vector of transmission of SARS-CoV-2^6,10^. This fact is especially because if infected they are mainly asymptomatic and have more difficulties in applying the correct procedures of the hygiene measures correctly^45,^. Interestingly, males reported higher relative frequencies than females in stating that children can get sick and transmit SARS-CoV-2. However, there’s no literature supporting this finding.

Regarding the use of gloves and/or masks, it is visible a certain hesitation and some myths associated with them, which may be explained by the quantity of contradictory information transmitted concerning these equipment^46^. The use of gloves is recommended during procedures associated with aerosol production, in a clinical context^46^. Regarding the global use of gloves, its general use is not recommended by WHO, CDC and DGS and may even constitute a false security if people don’t discard them regarding the proper procedure^47^. The use of masks has been under constant study since the beginning of this pandemic, in order to understand whether it prevents the infection by SARS-CoV-2 or not and the literature regarding this matter is very controversial. However, in a recent study, Greenhalgh *et al* advocate the general use of the mask, regardless of their material, even stating that a simple cotton mask will reduce the amount of transmitted virus by 36 times^48^. Indeed, masks are gaining a major role as a community preventive measure and not as an individual preventive measure^35,44,48,49,50^

Concluding, even though we observe a relatively well informed population, this is a pandemic of a virus with an elevated Transmission Rate. Thus, biological characteristics as well as individual and communitary behaviours have an impact in its course^10,14,20^ and even a single error/mistake may have a negative impact, creating new transmission chains that could have been prevented^32,33,43,44,62^. Following this thought, we are looking for the outliers of wrong answers/unfamiliarity of a concept and in order to promote a fruitful health literacy, towards an informed population and avoid an overwhelming unjustified panic, which may lead to more mistakes^51,52,53^.

To pursue this objective, we must take in account the major sources of information and adapt our communication, creating programs/methods in order to reach each specific population^54^. This adjustment to different realities has a positive impact on the population and their behaviour, by promoting community empowerment^55^.

On a final note, we safeguard that a correct health literacy does not always mean a correct behaviour and that some mistakes can still prevail^53,54^.

## 5. CONCLUSION

Health literacy is nowadays a critical issue and has an impact in controlling epidemics and pandemics^56,57,58^.

In Portugal we are now in phase 3.2 of mitigation of COVID-19’s pandemic and, even though we have a relatively well informed population, there seems to be some misconceptions of the guidelines^57^. As the actual public health situation is a pandemic by a highly infectious virus, every single deviant compliance matters and must be prevented. Thus, our goals were to characterize the population regarding their COVID-19’s health literacy, in order to help create specific intervention strategies aimed at populations with different levels of health literacy.

Although our cohort has a small dimension, it highlights some aspects that can bridge gaps that still remain. We believe it also provides support for future studies and alerts to the necessity of new approaches of communication in order to control not only this pandemic but also possible future pandemics with human-to-human transmission.

The main limitation of this work was the size of the database. Also, besides comparison between groups, other tests or applying metrics could have been used, e.g., correlation coefficients such as Spearman’s rank and Pearson’s. Also, a multinomial regression could have been carried out in order to predict the health literacy for an individual. Likewise, a new index variable representing health literacy could have been created, in order to state the level of health literacy of an individual. In further studies, this new variable can be created based on WHO and DGS information, enabling a comparative study of various coefficients, considering criteria and quality adjustment metrics.

## Data Availability

All the data collected was only accessed by the authors of this study.

## 6. DATA CONFIDENTIALITY

All data confidentiality was guaranteed.

